# The impact of physician-managed distribution of urological catheters on utilization

**DOI:** 10.1101/2022.08.11.22278049

**Authors:** Tom Hata, Jayme Hanson

**Author notes:** **Corresponding Author:** Tom Hata, PhD, 3040 VLSB #3140, Berkeley CA 94720. **Funding:** The authors received no specific public funding for this work.

## Abstract

**Introduction:** We examine whether provider-supplied urological catheters result in increased utilization by comparing claims data of providers before and after enrollment in a technology platform that allows them to directly order and manage distribution of prosthetics to patients.

**Methods:** We analyzed trends in per-provider quantity utilization of urological catheters by examining Medicare Part B claims data for HCPCS codes A4351, A4352, and A4353 (and an additional category, ALL CODES, which summed utilization across all 3 codes) from years 2014 to 2019. We then identified 64 referring providers who both submitted claims in at least one of the above three HCPCS codes in 2019 and transitioned to physician-managed distribution in 2021. Finally, we compared overall and per-beneficiary utilization by these providers between 2019 (traditional referral model) and 2021 (provider-supplied model) for each code category.

**Results:** We did not detect a significant increase in utilization for any code category. Overall utilization was not significantly different for code groups ALL CODES (p=0.26) and A4352 (p=0.8). Median A4351 utilization per provider decreased by 23% (p=0.01) after providers converted to the provider-supplied model. Correspondingly, median utilization of A4351 per beneficiary decreased by 23% (p=0.08) in the same span.

**Conclusions:** These findings show that provider-supplied catheter distribution to patients does not lead to increased utilization. In the case of HCPCS code A4351 catheters, physician-managed distribution may reduce wasteful oversupply of units to individual patients, resulting in an overall decrease in utilization.

## INTRODUCTION

Innovations in patient-centered care have driven many physician groups to provide integrated services to improve quality of care^1,2^, which includes an increase in self-referral of onsite services^3^. The advantages of integrated onsite services include enhanced patient/provider convenience^4^ and reduced handling errors^5^. However, there are concerns that self-referral creates a financial incentive for providers to increase utilization with the goal of financial gain^6,7^. To combat overutilization, congress passed Stark Law to prevent physicians from self-referring patients to designated health services (DHS) payable by Medicare or Medicaid. These services include but are not limited to, clinical laboratory services, radiology, home health services, outpatient prescription drugs, durable medical equipment, prosthetics, and orthotics supplies (DMEPOS)^8^. Although Stark Law has provided a means to protect federal health expenditures, in 2020, the Centers for Medicare and Medicaid Services (CMS) admitted that the laws have not kept pace to promote care coordination, improvement in quality or reduction in waste^9^.

There are numerous exceptions to Stark Law such as in-office ancillary services^8,10^, and studies of these exception cases have not universally demonstrated overutilization. Although some studies have observed increased utilization in certain contexts of self-referred imaging^11,12^ and testing^6,13^, there are a comparable number of studies that observed a lack of evidence of overutilization in others^3,14,15^. The mixed results of these studies in aggregate suggest that overutilization is not an inevitable consequence of physician-integrated services, and that risk for billing abuse by physicians should greatly factor into the determination of exceptions or clarifications to Stark Law.

Although Stark Law reform helped expand exceptions and clarify positions on value-based payment structures^16^, the law maintains prohibition of physician self-referrals for DHS, which includes services that support chronic patient management with high-rates of adverse events. Examples of the prohibited self-referred services are physician-provided supplies for the management of diabetes, urological bladder retention, bowel diversion, and wound care. However, the law does allow for physicians to bill for DMEPOS if they personally-perform the service^17^. A physician may be allowed to supply their patients directly as long as no other staff participates in the act of performing the service^17^. Prosthetics, such as catheters used for chronic urinary retention, potentially represent services that are high in medical certainty and low in potential for billing abuse by physicians. We examine the effects of physician-managed catheter supply on utilization by comparing claims from providers before and after they enrolled in services which enabled them to perform the duties of a DMEPOS supplier and provide prosthetics directly to their patients.

## METHODS

### Historical utilization in Medicare claims data

From the Medicare Durable Medical Equipment, Devices and Supplies by Referring Provider and Service dataset^18^, we selected claims data for HCPCS codes A4351, A4352, and A4353 (Appendix 1) for years 2014 to 2019 (the latest publicly-available data during the course of this study) to characterize utilization on a per-provider basis. We created an additional HCPCS category per provider, ALL CODES, which summed the utilization of the previous three codes per provider for each year to measure total unit utilization. We limited the scope of this study to quantity utilization, and we did not include monetary charge in our analysis. We identified regional and plan-based variation in charging rates as potential confounding factors. For each code category, we visualized data distribution and calculated median and total utilization per year.

**Appendix 1.** HCPCS codes used in study

| HCPSC Code | Description |
| --- | --- |
| A4351 | Intermittent urinary catheter; straight tip |
| A4352 | intermittent urinary catheter; coude (curved) tip |
| A4353 | Intermittent urinary catheter, with insertion supplies |

### Study population

We identified 64 referring providers who submitted claims in at least one of the above three HCPCS codes in 2019 and who have also enrolled and have submitted claims with Rx Redefined: a platform that allows physicians to directly order and manage the distribution of DMEPOS to their patients. We limited our scope of providers to those who were enrolled between 2019-2020 and were active users during 2021 to ensure a full year of use of service in 2021 (full-time enrollment of providers in 2020 was insufficient for robust quantitative analysis). We focused our comparison of utilization by these providers between their 2019 CMS and 2021 Rx Redefined claims submissions. The 2021 data of provider-supplied services include deidentified versions of both Medicare and commercial claims. We did not exclude commercial claims from analysis, because we chose not to assume that the proportion of Medicare beneficiaries per provider remained the same between 2019 and 2021. Rather, we decided to use the 2019 Medicare claims data as a conservative baseline for total utilization, given a lack of access to concurrent commercial claims data for these providers. We compared provider utilization between 2019 and 2021 for each code category using a two-sided Wilcoxon Signed Rank Test (a paired, non-parametric test).

Additionally, we compared utilization at a per-beneficiary level. Medicare Part B claims data suppresses the number of beneficiaries for a provider if the beneficiary count is below 11 for a given HCPCS code to protect patient privacy. Of the entries with beneficiary counts reported, we calculated average utilization per beneficiary of each HCPCS code for each provider. We excluded code categories A4353 and ALL CODES from further analysis, because there were no reported beneficiary numbers for A4353 in the Medicare dataset, and ALL CODES would not have provided any additional insight that could not be determined from comparing utilization of A43451 and A4352. We grouped and compared per-beneficiary utilization with the same treatment as overall utilization. All statistical analysis was done using Python 3.10 and the SciPy 1.8.0 package.

## RESULTS

Median utilization across every code category (fig. 1) displayed consistent uptrends (1 to 10% increases between successive years with no decreases) or no change (code A4353 years 2017-2019). Average values are represented as medians rather than arithmetic means because within a given year, utilization varied greatly amongst providers, spanning up to 4 orders of magnitude (10^1^–10^5^ units) and exhibiting a non-parametric, right-skewed distribution (fig. 2). Total utilization of each code category exhibited greater year-to-year variation (−7 to +18%) (fig. 3) than median utilization, likely due to year-to-year variance in provider utilization compounded by fluctuations in the number of referring providers in a given year (−8 to +14%) (fig. 4). Given relatively stable rates of utilization across each code category, we do not expect large year-to-year fluctuations to affect comparisons across years that are relatively close together. Of particular note, we expect changes in A4351 utilization to have the greatest effect on total utilization. A4351 represents >70% of total utilization in each year followed by A4352 (∼25%), and finally A4353 (∼3%) (fig. 3). Furthermore, A4351 was utilized by the greatest fraction of catheter-referring providers (>85% of providers, in contrast to ∼30% in A4352 and 12% in A4353) (fig. 4).

**Figure 1.**
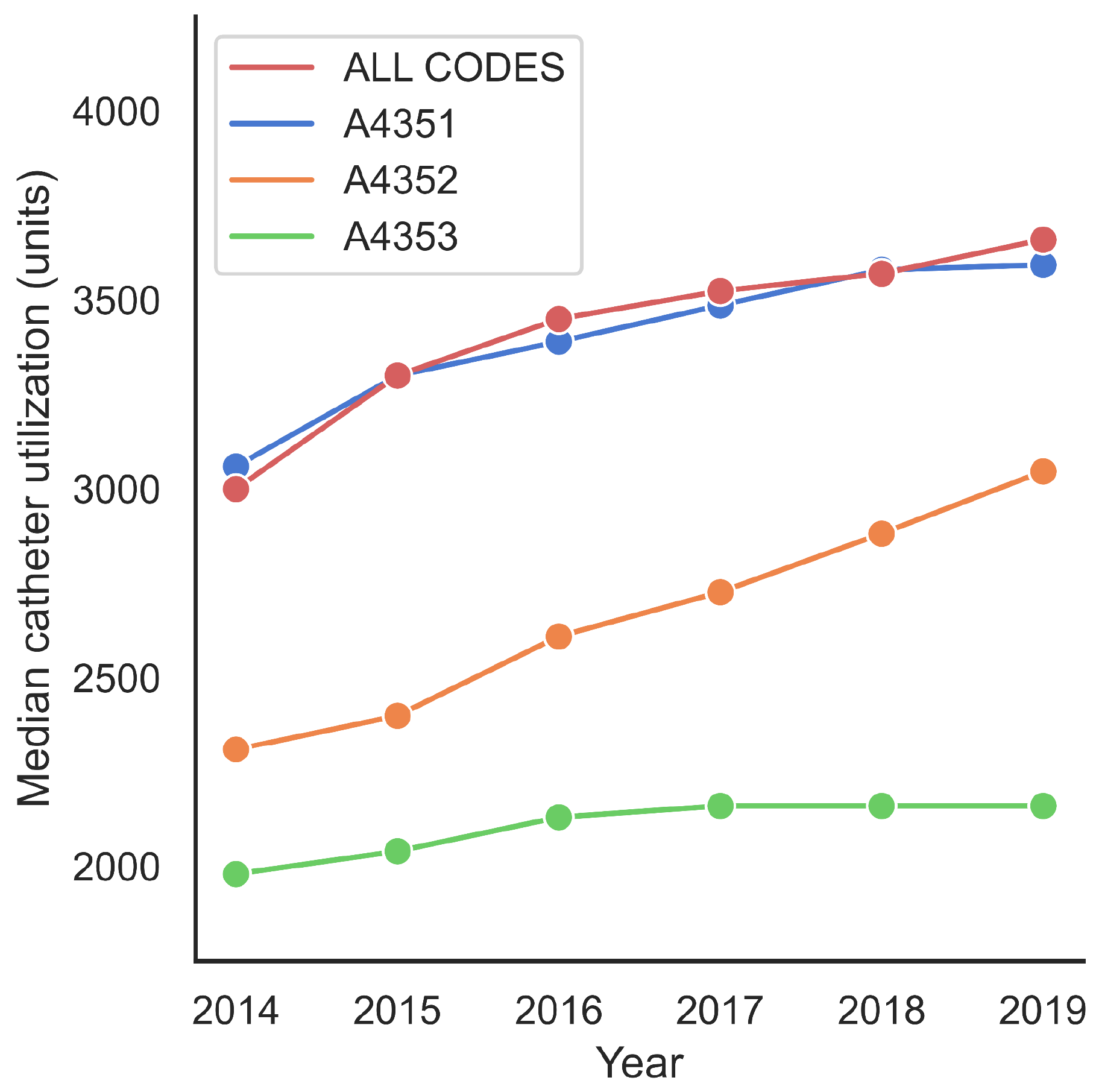
Median utilization of catheters per provider by HCPCS code from Medicare part B claims data.

**Figure 2.**
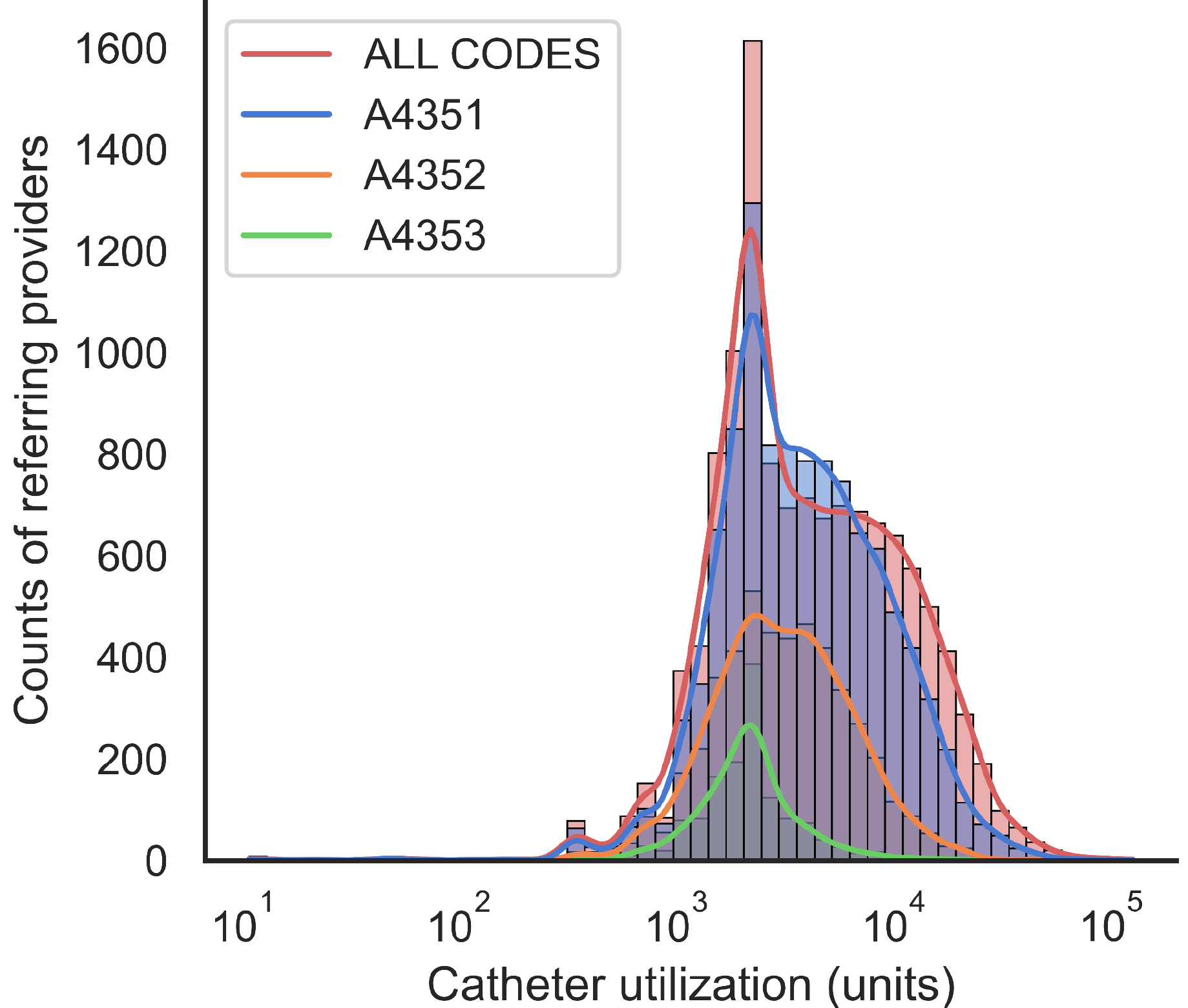
Histogram of referring provider counts (and kernel density estimate) binned by catheter utilization for 2019 Medicare part B claims. The distribution of data for 2019 is representative of other years observed in this study.

**Figure 3.**
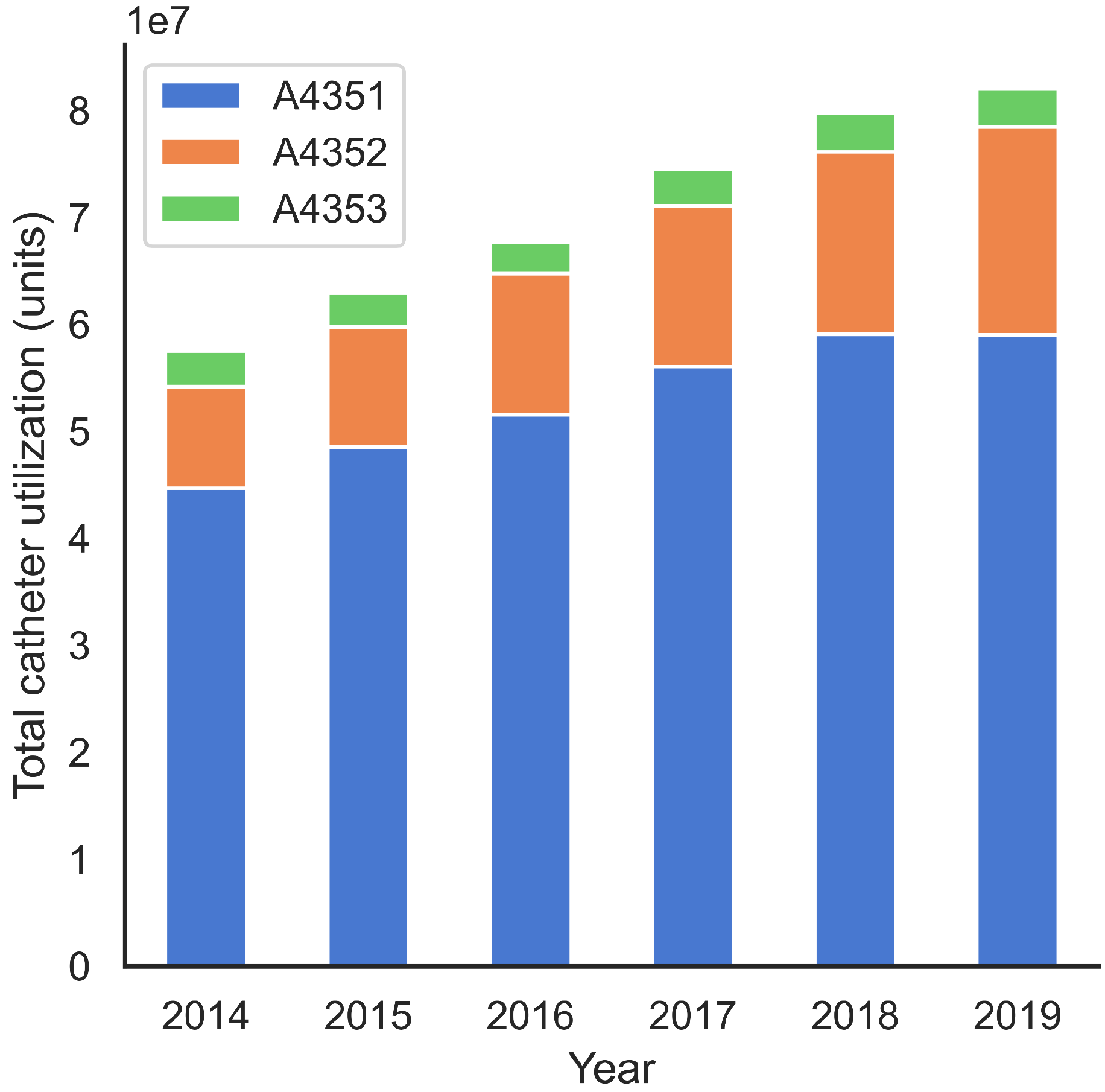
Total utilization of urological catheters by HCPCS code from Medicare part B claims data.

**Figure 4.**
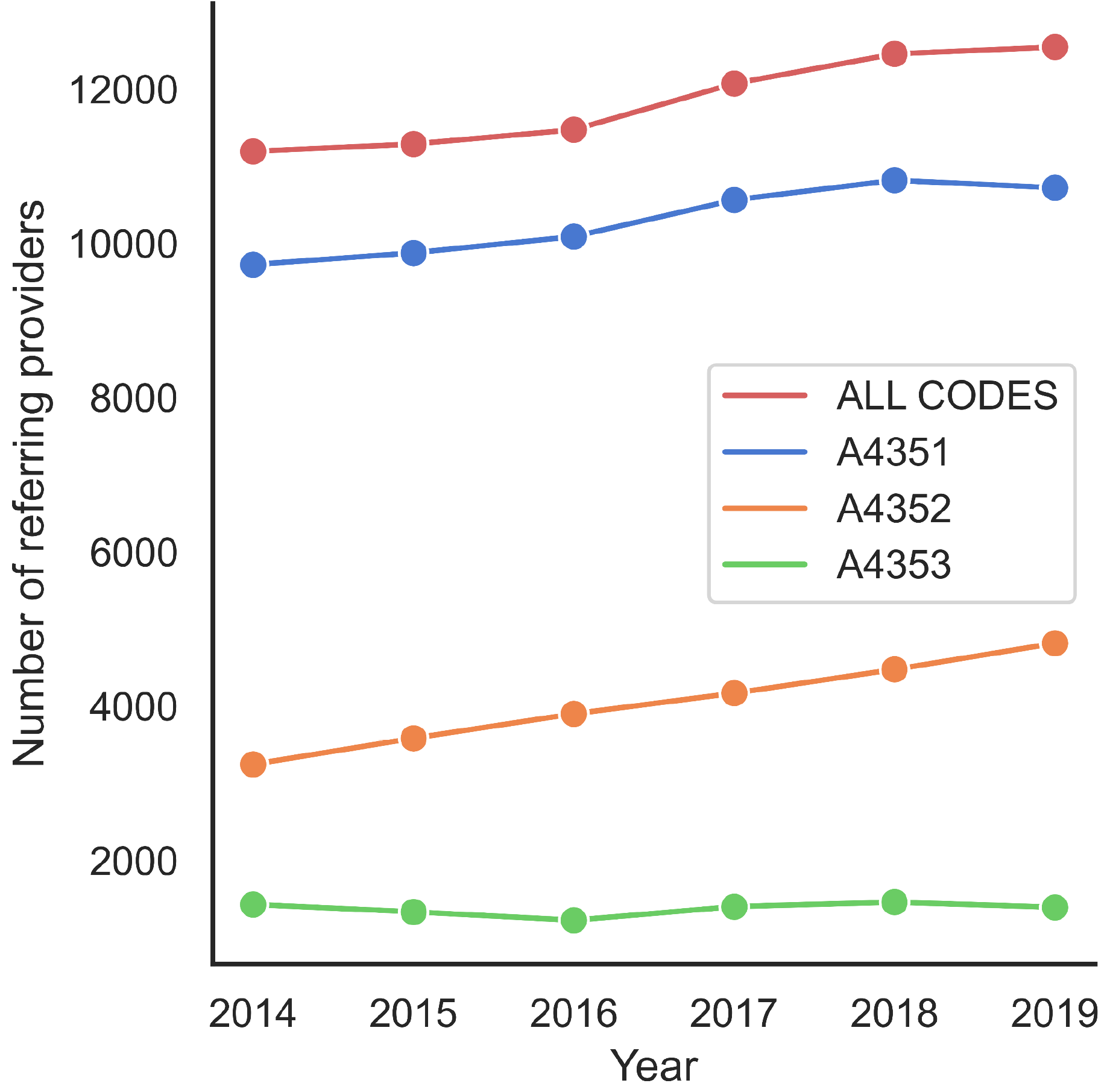
Number of referring providers who utilized catheter HCPCS codes A4351, A4352 and/or A4353 from Medicare part B claims data (a given provider may utilize one or more HCPCS code in a particular year).

We did not detect a statistically significant increase in utilization for any code category (fig. 5) when we compared 2019 and 2021 claims data. Physician utilization values between 2019 and 2021 were not statistically different for code groups ALL CODES (n=64, W=873, p=0.26) and A4352 (n=36, W=335, p=0.8) at the significance level of 0.05. In both cases, median utilization increased in 2021: from 7,788 to 8,505 units (+9%) in ALL CODES and from 3,896 to 4,155 units (+7%) in A4352. A4351 was the only code category to report a significant difference (n=61, W=598, p=0.01) in utilization between treatment years. Median utilization decreased in 2021, from 6,210 to 4,800 units (−23%). Statistical analysis was not conducted on code A4353 due to insufficient sample size (n=2), but median utilization did decrease 6,750 units to 4115 units (−39%).

**Figure 5.**
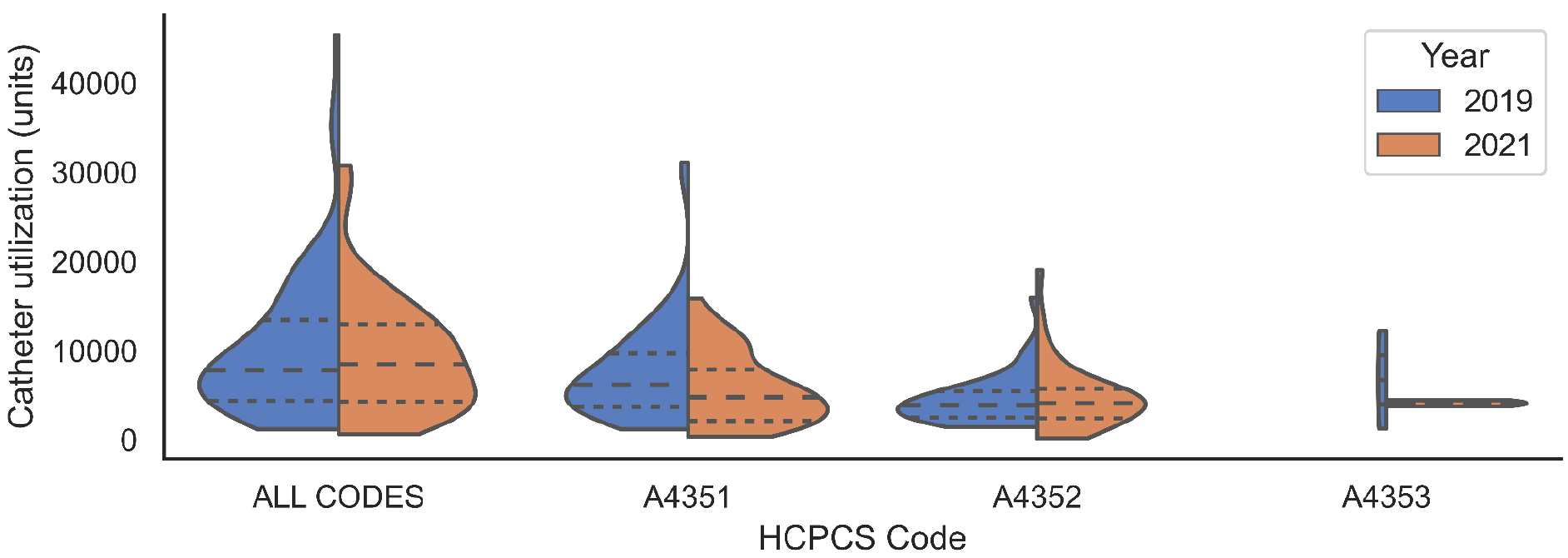
Violin plot of catheter utilization by referring providers, comparing 2019 Medicare part B claims against 2021 Medicare and commercial claims for physician-managed services. Dashed lines represent quartiles.

Median utilization per beneficiary decreased substantially for both codes A4351 (from 756 to 580 units: -23%) and A4352 (from 791 to 655 units: -17%) after providers converted from the traditional referral model in 2019 to the physician-managed model in 2021 (fig. 6). Although utilization rates per beneficiary were not statistically different at the significance level of 0.05 for either HCPCS code (A4351: n=20, W=57, p=0.08; A4352: n=4, W=3, p=0.6), A4351 utilization was statistically different at the significance level of 0.1.

**Figure 6.**
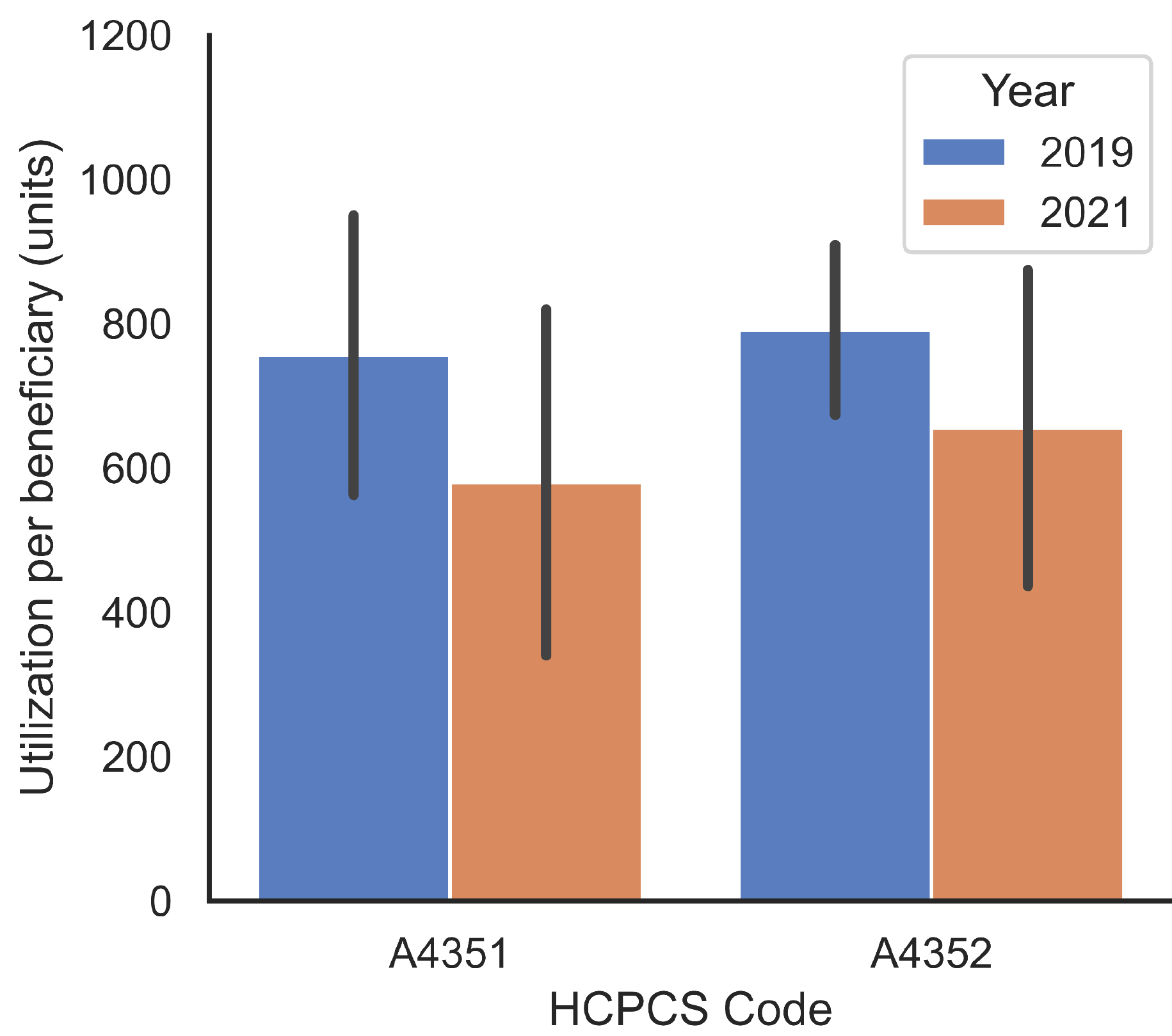
Median utilization per beneficiary (+/- 1 S.D.) for HCPCS codes A4351 and A4352, comparing 2019 Medicare part B claims against 2021 Medicare and commercial claims for physician-managed services.

## DISCUSSION

During preliminary analysis, utilization and charge to insurance were found to be tightly correlated to the point of redundancy, because the per-unit price did not vary greatly for a particular code. With limited to no control over price^19^, a provider seeking to abuse reimbursement claims for financial gain would therefore be driven to quantity overutilization. In our study population, utilization did not significantly increase after providers were given the means to directly supply DMEPOS to their patients (fig. 5). Increases in median utilization (A4352 and ALL CODES) fall in line with historical year-to-year variation found in the Medicare claims data. In contrast, the 23% decrease observed in median A4351 utilization may be primarily explained by a corresponding decrease of equal magnitude in per-beneficiary utilization (fig. 6). This may suggest that patients have already been overprescribed catheter units in the traditional referral model, and the observed decrease in utilization by physician-performed DHS could be explained in part by heightened ethical and oversight concerns of the service provider. A4351 units may be particularly incentivized by traditional distributors to be over-supplied, because they have the lowest rate of reimbursement^19^ and least amount of medical documentation required for prescription out of the three codes^20,21^.

Although the discussion of Medicare abuse is often centered around physicians as culprits, DMEPOS suppliers have proven to be significant sources of fraud^22,23^ in the traditional referral model. Downer^24^ found that of $1.7 billion in Medicaid fraud committed between 2014-2019 in the states of California, Florida, Georgia, New York, and Washington, $738 million (42%) of fraud was spent on DMEPOS suppliers. Relative to the demanding and highly-standardized path of becoming a physician, it is much easier and quicker for an individual to be granted a license to create a DMEPOS supply business. Therefore, a physician-integrated service model may reduce the prevalence of third-party bad actors inducing referral sources or starting DMEPOS businesses to take advantage of federal healthcare programs.

One caveat regarding the utilization data supplied by Rx Redefined is that total utilization may potentially be underestimated due to the nature of the service. After an initial ramp up period, the physician group attempts to handle all catheter DMEPOS claims whenever possible. However, patient enrollment is contingent on the provider offering and the patient accepting to use the physician-integrated supply services. As a result, some patients may still be guided through to a third-party channel to file their claims. Although our measurements of total utilization per provider may be impacted by this limitation, we do not expect calculations of per-beneficiary utilization rates to be affected.

## CONCLUSIONS

We believe this study shows that, at least in the context of urological catheters, provider-supplied DMEPOS services do not result in unethical behavior such as overutilization. Understanding that fraud is presently abundant in the third party / traditional referral pathway for DMEPOS, it may be sensible to promote the physician-integrated pathway for billable medical supplies where the barrier of entry, ethical expectations, and penalties for non-compliance are higher. Moreover, a physician-integrated model for chronic patients using DMEPOS is in support of recent adjustments in healthcare delivery and legislation to enable holistic care.

## Data Availability

The Medicare claims dataset is publicly available from Centers for Medicare & Medicaid Services. The RxRedefined claims dataset is propriety, and researchers interested in replication need to seek access by contacting RxRedefined.

https://data.cms.gov/provider-summary-by-type-of-service/medicare-durable-medical-equipment-devices-supplies/medicare-durable-medical-equipment-devices-supplies-by-referring-provider-and-service

## Notes

**Conflicts of Interest:** TH has received compensation from Rx Redefined as an independent researcher to analyze data and author this study. JH is employed by Rx Redefined.

### Competing Interest Statement

TH has received compensation from Rx Redefined as an independent researcher to analyze data and author this study. JH is employed by Rx Redefined.

### Author Declarations

The ethics committee of RxRedefined waived ethical approval for this study, as there was no risk to patients and we did not report identifiable data.

